# Optimal Pair Matching Combined with Machine Learning Predicts that Omega-3 Fatty Acid Supplementation Markedly Reduces the Risk of Myocardial Infarction in African Americans

**DOI:** 10.1101/2024.03.26.24304910

**Authors:** Shudong Sun, Aki Hara, Laurel Johnstone, Brian Hallmark, Eric Lu, Joseph C. Watkins, Cynthia A. Thomson, Susan M. Schembre, Susan Sergeant, Jason Umans, Guang Yao, Hao Helen Zhang, Floyd H. Chilton

## Abstract

Conflicting results from clinical trials have contributed to a lack of consensus about cardioprotective effects of omega-3 (n-3) highly unsaturated fatty acids (HUFA). Although the VITAL trial did not demonstrate an overall benefit of n-3 HUFA supplementation on composite cardiovascular disease (CVD) and cancer outcomes, the substantial enrollment of African Americans (AfAm) afforded a unique opportunity for a post-hoc analysis of racial differences in the supplementation response. We employed propensity score matching to address potential covariate imbalances between AfAm and European American (EuAm) participants in VITAL (N=3,766 participants). Using Kaplan-Meier curves and two machine learning methodologies, we found that n-3 HUFA supplementation was significantly associated with a reduced risk of myocardial infarction (MI) exclusively in the AfAm subgroup, as evidenced by an odds ratio of 0.17 (95% CI [0.048, 0.59]). These findings indicate a potential cardioprotective benefit of n-3 supplementation in AfAm, specifically in reducing MI risks; a risk not identified in EuAm. Further investigation of n-3 HUFA effects through a hypothesis-driven randomized clinical trial among AfAm is needed to test a race-specific response that may inform recommendations for n-3 HUFA supplementation.

## Introduction

In 1999, the landmark GISSI-Prevenzione (GISSI-P) randomized, placebo-controlled trial (RCT) was published reporting the effects of daily supplementation with 1 g/day of n-3 highly unsaturated fatty acids (n-3 HUFAs), eicosapentaenoic acid (EPA) + docosahexaenoic acid (DHA), in patients (N=2,836 n-3 HUFA vs 2,830 placebo) who had recently experienced a myocardial infarction (MI).^1^ Over the 4-year study, researchers reported a 14% risk reduction in the combined primary endpoint of relative risk and an overall 20% reduction in fatal cardiovascular events. This groundbreaking study, however, was met with mixed results in subsequent studies, which questioned the additive benefits of n-3 HUFA supplementation to modern cardiovascular therapies, particularly at lower doses^.2,3^ Further investigations, including a 2018 comprehensive systematic review, containing 79 RCTs with 112,059 patients found an overall 8% reduction in cardiac death associated with n-3 HUFA supplementation.^4^

Yet, the landscape of n-3 HUFA research continues to be marked by variability, as seen in the differing outcomes of notable recent large RCTs such as ACEND, VITAL and REDUCE-IT. ACEND examined the impact of n-3 HUFA supplementation on individuals with diabetes, finding no significant differences in the risk of serious vascular events between n-3 HUFA supplementation and placebo groups.^5^ VITAL tested the impact of n-3 HUFA and vitamin D3 supplementation on primary prevention of cardiovascular disease (CVD) and cancer, again finding no difference between the n-3 HUFA intervention and placebo groups. In contrast, the REDUCE-IT clinical trial provided higher doses of EPA as an ethyl ester to patients at high risk of CVD and demonstrated a 25% reduction in composite CVD morbidity and mortality (including CVD mortality, non-fatal MI, non-fatal stroke, cardiovascular revascularization or unstable angina) in the treatment group^6^. Subsequent meta-analyses and reviews suggest discordant clinical trial results are likely related to numerous factors, including different doses and forms of n-3 HUFAs provided, background dietary levels of n-3 HUFAs, CVD and diabetes-related risk factors, and medications, each of which may limit the measurable effects of n-3 HUFAs^.7–10^

The RCTs to date have generally demonstrated a lack of diversity across the study populations, implicitly suggesting a one-size-fits-all response to n-3 HUFA supplementation across diverse racial groups. Yet, arachidonic acid (ARA) production and metabolism, a major drivers of CVD risk,^11^ varies widely across racial groups related to differences in the frequency of key genetic variants particularly within the fatty acid desaturase (*FADS*) gene cluster that influence the biosynthesis of HUFAs. Importantly, compared to EuAm, AfAm exhibit significantly higher frequencies of fatty acid desaturase (*FADS)* genetic variants which are linked to a more efficient conversion of the dietary omega-6 (n-6) polyunsaturated fatty acid (PUFA) linoleic acid (LA) to ARA^12,13^. These genetic factors and other race-associated variation^12^ coupled with the high dietary levels (6-8% total energy intake) of LA in Western diets (MWD) have the potential to contribute to pronounced differences in the balance of ARA-derived pro-inflammatory and pro-thrombotic metabolites relative to EPA and DHA-based anti-inflammatory, anti-thrombotic, and pro-resolution oxylipin profiles among AfAm compared to EuAm populations.^14^

Capitalizing on the diversity of participants in the VITAL trial, with its substantial enrollment of AfAm participants (N= 5,106), Manson et al. in a follow-up analysis of VITAL, examined potential modifiers of the n-3 HUFA treatment effects including race and ethnicity and showed that AfAm experienced a significant 77% treatment-associated reduction in MI (HR=0.23 [0.11– 0.47]) while other racial and ethnic groups had smaller reductions (p, interaction=0.001^15^). Similarly, a follow-up Kaplan-Meier analysis of the VITAL data by our group demonstrated a marked ∼80% reduction in MI associated with n-3 HUFA supplementation in AfAm participants with baseline cardiovascular risk who did not consume fish^.16^ A recent analysis of VITAL using hierarchical composite CVD outcomes based on win ratio demonstrated that participants with low fish consumption at baseline benefited more than those with high consumption when randomized to receive n-3 HUFA supplementation^.17^ Despite the consistency of these findings, there are limitations of *post-hoc* analyses that must be addressed to ensure the reliability of such promising findings.

The goal of the current study was to employ propensity score matching to further address limitations of secondary data analysis related to confounding variables. Specifically, we paired EuAm participants with an AfAm subgroup, based on their covariate profiles and then leveraged machine learning methods to carefully curate a dataset of 3,766 AfAm and EuAm individuals. Using this methodology, we were able to simulate RCT conditions within observational data to assess risk of MI, stroke and CVD mortality with n-3 HUFA supplementation in the AfAm group as compared to EuAm. Findings from this study provide the best evidence to date that highlight the potential of n-3 HUFA supplementation to prevent MI in AfAm individuals. They also emphasize the necessity for considering race in the design and power calculations of future trials utilizing n-3 HUFA supplementation.

## Results

### Utilization of Optimal Pair Matching to Balance Potential Confounding Variables

The original VITAL trial had n=25,871 participants with n=18,046 (71%) EuAm, n=5,106 (20%) AfAm. The remaining 9% belong to other racial or ethnic groups. This study considered 19 variables in EuAm and AfAm groups. After exclusions (see Methods), data from n=19,319 participants (n=15,553 EuAm and n=3,766 AfAm) were utilized to compare the preventive effects of n-3 HUFA supplementation against MI, stroke, and CVD mortality. As noted earlier, the original VITAL study did not stratify based on race to compare these groups and many covariates had markedly different distributions across racial groups (**Supplementary Table 1**). For example, the average age for EuAm was 67.4 years vs 62.4 years for AfAm. The smoking rate was 14% in EuAm vs 5% in AfAm, and the diabetes comorbidity was 23.1% in EuAm and 10% in AfAm.

To reduce bias arising from confounding variables, propensity score-based matching was employed. Three one-to-one matching methods were explored: 1) optimal pair matching^18,19^, 2) nearest neighbor matching^20,21^, and 3) genetic matching^22^. Optimal pair matching was selected for all subsequent analyses due to its superiority in minimizing the standardized mean difference between matched pairs (see Methods). The characteristics of the pared-down cohort (n=3,766) AfAm and EuAm participants after optimal pair matching are detailed in **Table 1**. Important variables such as age, smoking, medications, fish consumption, and cardiovascular risk factors were comparable between AfAm and EuAm participants in this carefully matched dataset.

**Table 1:** Demographic Table after Optimal Pairs Matching.

| Characteristic |  | Total<br>(N=7532) | Af Am<br>(N=3766) | Eu Am<br>(N=3766) |
| --- | --- | --- | --- | --- |
| Age |  | 62.8±6.3 | 62.4±6.6 | 63.1±5.9 |
| Female |  | 4587 (60.9) | 2347 (62.3) | 2240 (59.5) |
| BMI |  | 30.3±6.6 | 30.6±6.5 | 29.9±6.1 |
| Current smoking |  | 979 (13.0) | 528 (14.0) | 451 (12.0) |
| Hypertension medication |  | 4754 (63.1) | 2446 (64.9) | 2308 (61.3) |
| Cholesterol medication |  | 2451 (32.5) | 1200 (31.9) | 1251 (33.2) |
| Diabetes |  | 1617 (21.5) | 871 (23.1) | 746 (19.8) |
| Diabetes medication |  | 1267 (16.8) | 675 (17.9) | 592 (15.7) |
| Parental history of MI |  | 1161 (15.4) | 591 (15.7) | 570 (15.1) |
| Fish consumption (≥1.5/wk) |  | 3750 (49.8) | 1890 (50.2) | 1860 (49.4) |
| Aspirin use |  | 2885 (38.3) | 1431 (38.0) | 1454 (38.6) |
| Statin use |  | 2279 (30.3) | 1114 (29.6) | 1165 (30.9) |
| Vitamin D supplements |  | 2271 (30.2) | 1096 (29.1) | 1175 (31.2) |
| CVD risk factors | 0 | 1661 (22.1) | 778 (20.7) | 883 (23.4) |
|  | 1 | 2634 (35.0) | 1315 (34.9) | 1319 (35.0) |
|  | >1 | 3237 (43.0) | 1673 (44.4) | 1564 (41.5) |

### Effect of n-3 HUFA Supplementation on MI in AfAm and EuAm Participants

Following this matching approach, Kaplan-Meier analysis was initially used to compare rates of MI between groups receiving n-3 HUFA supplementation and those given placebo, stratified by race. **Figure 1** illustrates the resulting survival curves and MI events over the six-year study. AfAm participants who were randomized to n-3 HUFA supplementation showed a marked reduction in MI compared to those who received placebo (p=0.00034), indicating a statistically significant effect in this subgroup. Conversely, no significant reduction in MI was observed in EuAm participants (p=0.59).

**Figure 1:**
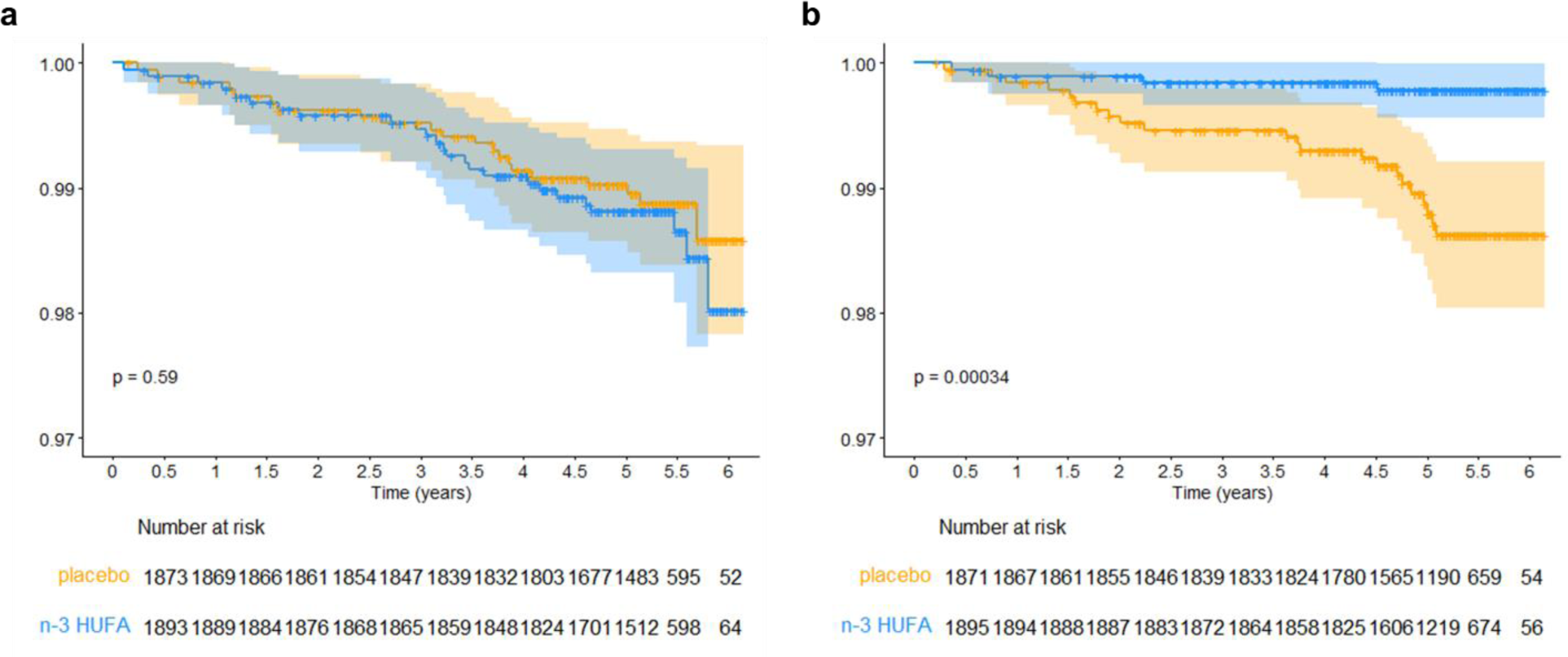
Kaplan-Meier Survival Curves for Myocardial Infarction in n-3 HUFA Treatment and Placebo Groups, Faceted by Race/Ancestry, for Participants Selected by Optimal Pair Matching. a. Kaplan-Meier plots and the number of subjects at each event point for EuAm participants matched with AfAm participants. b. Kaplan-Meier plots and the number of subjects at each event point for AfAm participants. n-3 HUFA, n-3 HUFA supplementation group; placebo, placebo group. The tables below the plots indicate the number of subjects in each group still in the study and MI-free at each six-month time point. Unadjusted p-values for the treatment effect were calculated for each racial group using the log-rank test.

### Logistic Regression Analysis with LASSO to Select Important Variables and Bootstrap to Estimate the Standard Errors

Logistic regression analysis with interaction terms between n-3 HUFA supplementation and other variables (including race), employing the Least Absolute Shrinkage and Selection Operator (LASSO) for variable selection and bootstrap methods for estimating standard errors, identified n-3 HUFA supplementation as the most significant predictor of MI incidence among AfAm participants (**Table 2**). These series of analyses were performed on a combined population of matched AfAm and EuAm, and race was factored in as a variable. The odds ratio (OR) for MI incidence with n-3 HUFA supplementation compared to placebo was 0.17, with a 95% confidence interval (CI) ranging from 0.048 to 0.60. Conversely, for EuAm participants, the analysis indicated no significant effect of n-3 HUFA supplementation on MI risk, with an OR of 1.0 and a 95% CI of [0.74, 1.34]. These findings remained consistent across both non-parametric and parametric bootstrap analyses (**Table 2**).

**Table 2:** Regression Results after Selection of Variables by LASSO with MI as the Outcome Using African American and European American Participants after Optimal Pairs Matching. Includes interaction terms. Std. Errors, p-values and 95% CI for OR are calculated using non-parametric bootstrap and parametric bootstrap. OR, odds ratio; CI, confidence interval. n-3 HUFA supplementation OR (95% CI) for AfAm and EuAm subgroups.

| Variables | Estimate | OR | Non-parametric Bootstrap |  |  | Parametric Bootstrap |  |  |
| --- | --- | --- | --- | --- | --- | --- | --- | --- |
|  |  |  | Std. Error | P-value | 95% CI for OR | Std. Error | P-value | 95% CI for OR |
| (Intercept) | -3.7599 | 0.0233 | 1.9965 | 0.0597 | (0.0005, 1.1656) | 1.8195 | 0.0388 | (0.0007, 0.8239) |
| Female | -0.7184 | 0.4875 | 0.3501 | 0.0402 | (0.2455, 0.9683) | 0.3448 | 0.0372 | (0.2480, 0.9583) |
| Age | 0.0552 | 1.0568 | 0.0277 | 0.0463 | (1.0009, 1.1157) | 0.0251 | 0.0279 | (1.0060, 1.1100) |
| BMI | 0.0304 | 1.0309 | 0.0249 | 0.2221 | (0.9818, 1.0824) | 0.0234 | 0.1939 | (0.9847, 1.0792) |
| Current Smoker | 0.7167 | 2.0477 | 0.4155 | 0.0845 | (0.9069, 4.6232) | 0.4471 | 0.1089 | (0.8525, 4.9186) |
| Diabetes | 0.4599 | 1.5839 | 0.4255 | 0.2798 | (0.6879, 3.6469) | 0.3381 | 0.1738 | (0.8165, 3.0728) |
| Fish consumption (1.5/wk) | -0.7424 | 0.4760 | 0.3755 | 0.0480 | (0.2280, 0.9936) | 0.3274 | 0.0234 | (0.2505, 0.9042) |
| n-3 HUFA supplementation x Af Am | -1.7747 | 0.1695 | 0.6410 | 0.0056 | (0.0483, 0.5955)* | 0.6944 | 0.0106 | (0.0435, 0.6613)* |
| n-3 HUFA supplementation x Eu Am | 0.0000 | 1.0000 | 0.1508 | 1.0000 | (0.7442, 1.3438)* | 0.1566 | 1.0000 | (0.7357, 1.3593)* |

### Weighted Decision Tree Analysis of MI in AfAm and EuAm Participants

Weighted decision trees were constructed to identify the most predictive variables of MI incidence among AfAm and EuAm group, analyzed separately. **Figure 2a** presents the decision tree for AfAm participants, where the first branching point underscores that assignment to the n-3 HUFA supplementation group as the predominant factor for MI prevention in this subgroup. Subsequent branches identify BMI, age, and the presence of diabetes as significant variables influencing MI incidence among AfAm participants. Conversely, the decision tree for EuAm participants, shown in **Figure 2b**, shows that n-3 HUFA supplementation did not significantly affect MI incidence. Instead, BMI and sex emerged as primary variables associated with MI outcomes.

**Figure 2:**
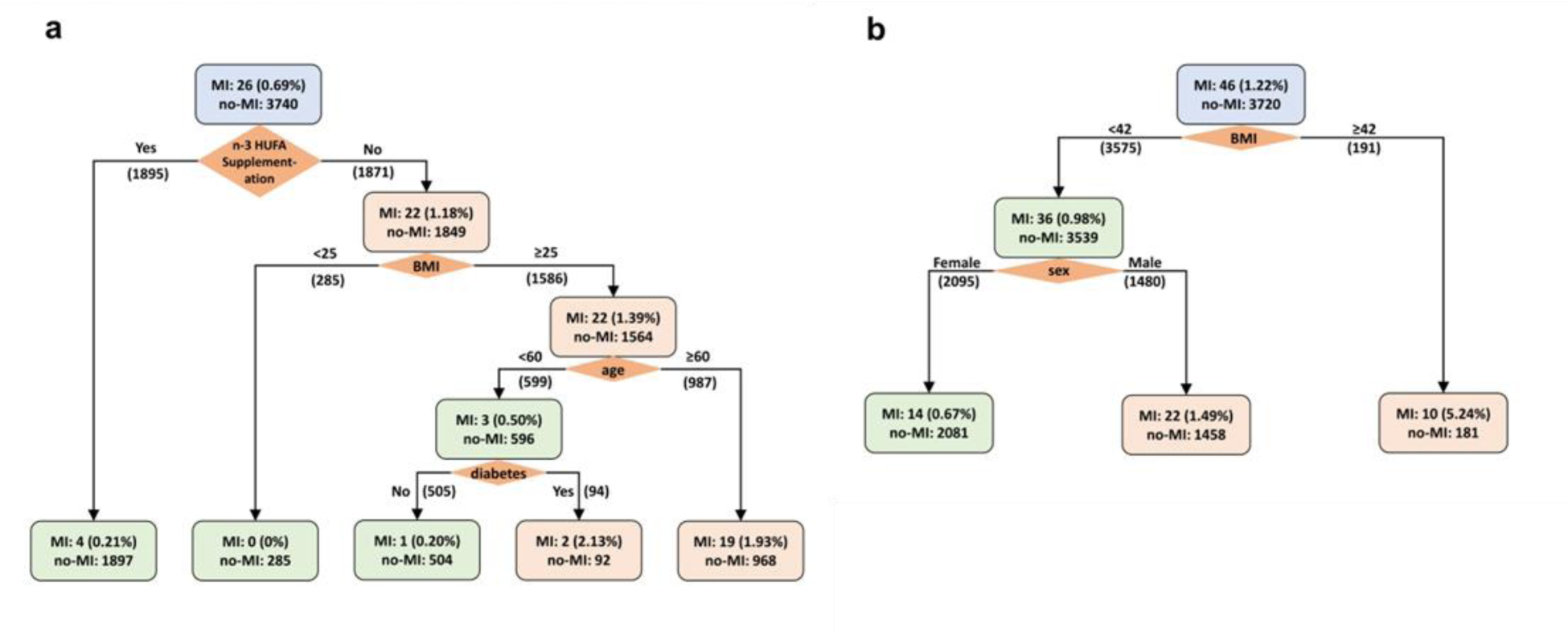
Weighted Decision Tree for Predicting the Incidence of Myocardial Infarction. a. Decision Tree for AfAm participants after optimal pair matching, with a total of n=3,766. The weight ratio of non-diseased to diseased participants is 3,740:26, equating to 144:1. b. Decision Tree for EuAm participants after optimal pair matching, with a total of n=3,766. The weight ratio of non-diseased to diseased participants is 3,720:46, which corresponds to 81:1. n-3 HUFA supplementation, active or placebo; BMI, body mass index at randomization, kg/m2; age, age at randomization to VITAL study, years; diabetes, baseline diabetes, yes or no; sex, female or male.

### Weighted Decision Tree and Lasso Regression Analysis of Stroke or CVD Mortality in AfAm and EuAm Participants

Neither the decision tree nor the logistic regression with LASSO analyses identified randomization to n-3 HUFA supplementation as an important factor in influencing stroke or CVD mortality in AfAm or EuAm groups (**Figure 3, Supplementary Figure 2, Supplementary Tables 2 & 3**). The decision tree for stroke identified age and BMI as predictive factors for both AfAm and EuAm participants. Additionally, statin use was found to be a significant factor within the AfAm subset (**Figure 3**). Where the outcome was CVD mortality, age, aspirin use, and BMI were significant factors for the AfAm subset, whereas for EuAm participants, predictive factors were limited to current smoking and BMI (**Supplementary Figure 2**).

**Figure 3:**
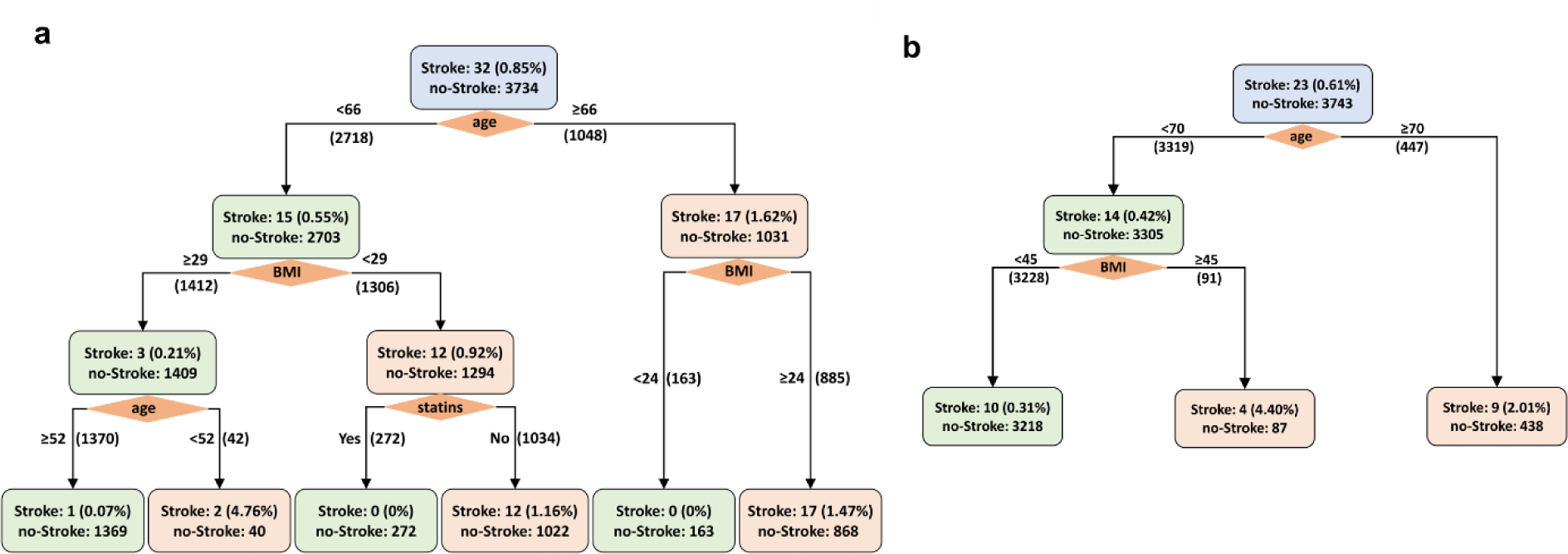
Weighted Decision Tree for Predicting the Incidence of Stroke. a. Tree for AfrAm participants after optimal pairs matching, n=3,766, weight ratio = non-diseased: diseased= 3734:32 = 117:1. b. Tree for EuAm participants after optimal pair matching, n=3,766, weight ratio = non-diseased: diseased = 3743:23 = 163:1. BMI, body mass index at randomization, kg/m2; age, age at randomization to VITAL study, years; statins, baseline statin use, yes or no; sex, female or male.

In the logistic regression model addressing stroke as the outcome, only age remained a significant factor. Although assignment to the n-3 HUFA group was among the variables selected by the LASSO algorithm, its interaction with race did not prove to be significant for either the AfAm or EuAm subgroups (**Supplementary Table 2**). Similarly, in the regression analysis focusing on CVD mortality, age and current smoking were the only significant predictors, and n-3 HUFA supplementation did not appear among the LASSO-selected variables (**Supplementary Table 3**). Both regression models were further validated using parametric bootstrap analysis, yielding results consistent with those obtained from the non-parametric bootstrap approach (**Supplementary Tables 2 & 3**)

## Discussion

In analysis of the comprehensive data on CVD endpoints and covariates from the VITAL trial, our findings confirm there was a striking reduction (OR estimate 0.17, CI [0.048, 0.60]) in MI among AfAm individuals as a result of randomization to n-3 HUFA supplementation versus placebo. This finding may be of fundamental importance, considering the disproportionately elevated rates of MI incidence and worse outcomes within this demographic^.23^ While the VITAL trial’s overall analysis did not show a significant reduction in major cardiovascular events across racial groups, our targeted analyses revealed potential benefits of n-3 HUFA supplementation may vary for AfAm as compared to EuAm. This outcome aligns with previous secondary analyses conducted by Manson et al.^15^and our group,^16^ and suggesting that aggregating data across racial groups may obscure critical findings related to race-specific effects. Importantly, these data provide evidence that n-3 HUFA supplementation could play a key role in addressing the elevated MI risk among AfAm populations.

Recognizing the initial disparities in covariate distributions between AfAm and EuAm participants, our study initially leveraged propensity score matching to achieve a more balanced and equitable comparison. This strategic approach not only refined the EuAm sample for parity but also allowed us to simulate a RCT scenario that compensated for the differences in baseline characteristics. This, in turn, enabled a more reliable assessment of the effect of randomization to n-3 HUFA supplementation versus placebo for AfAm and EurAm subgroups. These compelling findings underscore the urgent need for a deeper exploration into whether n-3 HUFA supplementation differentially influences health outcomes across racial (and ethnic) subgroups. A well-designed, sufficiently powered randomized clinical trial focused exclusively on evaluating the hypothesis that n-3 HUFA supplementation has a pronounced effect on reducing MI risk among AfAm population is warranted. Such investigations could yield crucial insights into mechanistic drivers of n-3 supplementation effects by race and inform on future precision approaches to cardiovascular care, ultimately facilitating the development of more effective prevention strategies to modify MI risk.

There are known molecular and genetically influenced mechanisms that may explain why n-3 HUFAs, and particularly EPA-enriched supplementation reduces the risk of MI in AfAm but not EuAm. Numerous studies have established that the efficiency of the HUFA biosynthetic pathway is under genetic control, with variations in the *FADS* gene cluster exerting the most significant influence on n-6 and n-3 HUFA levels^.24^ The link between *FADS* variants and complex lipid and inflammatory phenotypes is well-documented. In fact, the *FADS1-FADS2* gene region has been identified as a critical multimorbidity-associated cluster in the human genome.^25^ *FADS* associations have been confirmed in both EuAm and AfAm cohorts, with a meta-analysis involving approximately 8,000 AfAm participants validating the connection between *FADS* single nucleotide polymorphisms (SNPs) and lipid phenotypes as well as coronary artery disease.^26^

Further, the frequencies of *FADS* genetic variants that affect HUFA biosynthesis, especially ARA, vary significantly across human populations^27^. Although self-reported race does not equate to genetic ancestry, there is a strong correlation in the context of *FADS* variation^.12,13, 27^ Approximately 80% of AfAm have homozygous ‘derived’ alleles linked to efficient HUFA metabolism, in contrast to about 43% of EuAm individuals. At the population level, these genetic and other factors significantly alter the balance between the n-6 HUFA, ARA and n-3 HUFAs, such EPA and DHA^12,13^. Importantly, epidemiology studies highlight a strong association between higher EPA/ARA ratios and a reduced risk of future CVD events.^28,29^

If the ratio of ARA to EPA or DHA indeed reflects the synthesis of n-6 and n-3 HUFA-derived oxylipins, as supported by research,^30^ then AfAm may experience a significant imbalance towards pro-inflammatory and pro-thrombotic versus anti-inflammatory/anti-thrombotic/pro-resolutive oxylipins leading to MI^.30,31^ The serious consequences of such an imbalance were underscored two decades ago with the removal of selective cyclooxygenase 2 inhibitors from the market, following cardiovascular events driven by a detrimental shift in the balance between pro-thrombotic thromboxane and anti-thrombotic prostacyclin.^32,33^ Collectively, this leads to the hypothesis that the levels and ratios of dietary n-6/n-3 PUFAs-influenced by the Western diet ^34^ result in a marked imbalance in ARA versus EPA and DHA, along with their oxylipin derivatives. This imbalance is notably exacerbated in AfAm populations where the genetic variation in *FADS* and other race-associated factors substantially enhances the capacity for ARA production^,12–14^ thereby tipping the balance toward ARA and its downstream pro-inflammatory and prothrombotic oxylipins, and away from EPA and DHA with their anti-inflammatory and anti-thrombotic effects. As a result, supplementation with n-3 HUFAs, which are known to balance the ratio of ARA to EPA and enhance EPA-derived oxylipins^,35^ could be especially beneficial for AfAm populations, where the extent of this imbalance is so pronounced.

The primary limitation of this study is its post-hoc nature. The VITAL study was not initially designed to examine racial differences in response to n-3 HUFA supplementation versus placebo. Despite our efforts to address limitations of post-hoc analyses using propensity score matching, these findings should be interpreted with caution, as they do not directly infer causality. Moreover, while propensity score matching attempts to minimize confounding in observational studies, it relies on the assumption that all relevant variables have been measured and included in the model. This method also reduces sample size, potentially decreasing statistical power and increasing the risk of Type II errors. Despite this, findings from this study underscore the importance of further investigating racial differences in MI risk related to n-3 HUFA supplementation. These findings enhance our understanding of MI health disparities in AfAm and emphasize the need for more focused hypothesis-driven research to establish causal relationships between n-3 HUFA supplementation and health in AfAm.

## Methods

### Data Exclusion

Variables with more than 20% missing data were excluded. For the remaining variables, only complete cases (those where a subject has no missing data) were retained.

### Propensity Score Matching

Propensity score matching^20,21^ was employed to select matched EuAm individuals for each AfAm individual in the dataset. The input data for matching comprised the dataset with missing values processed. The R package *MatchIt*^36^ was utilized to implement three different matching methods,^18–22^ optimal matching, nearest neighbor matching, and genetic matching. The first two methods are based on estimating the propensity score using logistic regression. Nearest neighbor matching, also known as greedy matching, involves sequentially pairing treated units with the closest eligible control unit. Optimal pair matching aims to select matches that collectively optimize an overall criterion. In our case, the criterion was the sum of absolute pair distances in the matched sample. The third method, genetic matching, employed a genetic algorithm, which optimized non-differentiable objective functions, to determine scaling factors for each variable in a generalized Mahalanobis distance formula. The algorithm seeks to achieve covariate balance. After obtaining the scaling factors, nearest neighbor matching was performed using the scaled generalized Mahalanobis distance. To evaluate the performance of different matching methods, the standardized mean difference (SMD) was evaluated, and optimal pair matching outperformed the other approaches.

### Survival Analysis and Kaplan-Meier Curves

Subgroup analysis was performed using the VITAL trial data for the outcome of MI, stratified by race. Only EuAm and AfAm subjects selected by the optimal pairs matching algorithm were included. Statistical calculations were performed in R (version 4.2.2) using the surviva^l37^ (version 3.4-0) and survminer (https://rpkgs.datanovia.com/survminer/index.html, version 0.4.9) packages. Kaplan-Meier survival curves were produced separately for each racial group, and p-values for the effect of the n-3 HUFA treatment on the occurrence of MI were calculated using the log-rank test.

### Logistic Regression Analysis with LASSO for Variable Selection and Bootstrap for Standard Error Estimation

For this analysis, both racial subgroups were included in one dataset with race encoded as a variable along with all other variables retained after filtering for missing data (see Data Exclusion above). Interaction terms between n-3 HUFA supplementation and all other variables were introduced to identify potential interactions with n-3 HUFA supplementation. To classify patients with MI (or stroke or cardiovascular death) versus those without MI, a weighted LASSO regression model^38^ was built using the *glmnet*^39^ package in R for variable selection. A 5-fold cross-validation method was employed to tune the LASSO model. Subsequently, a logistic regression model was fitted considering only the variables selected by the LASSO model. To estimate the standard errors, both non-parametric bootstrap and parametric bootstrap methods^40^ were employed. Using the logistic regression coefficients and the standard errors estimated through bootstrapping, p-values, odds ratios, and corresponding 95% confidence intervals were calculated. During the bootstrap procedure, models with large distances for critical coefficients were excluded to avoid drawing poor samples.

### Decision Tree

The matched data were utilized as input to build a predictive model, specifically a decision tree, using the classification and regression trees (CART) algorithm^41^ implemented in the R package *rpart.* ^42^ The AfAm and EuAm subgroups were independently utilized to construct the trees, facilitating a comparison of the outcomes. Since the dataset was severely unbalanced, a weighted tree algorithm was applied with the weight ratio equal to the number of individuals with MI (or stroke or cardiovascular death) to the number of individuals without these outcomes. The Gini index was employed to determine the tree split points, and a minimum node size of 20 was set as a requirement. To optimize the tree model and assess its prediction accuracy, a 5-fold cross-validation method was employed. To prevent overfitting, the tree was pruned using the 1-SE rule. This rule identifies the number of splits that yield the smallest cross-validation error, adds the corresponding standard error, and selects the fewest splits where the cross-validation error remains smaller than this combined value.

## Data Availability

All data produced in the present study are available upon reasonable request to the authors
All data produced in the present work are contained in the manuscript
All data produced are available online at https://data.projectdatasphere.org/projectdatasphere/html/content/454

---

**Supplementary Table 1:** Demographic Table before Matching (Original Participant Set, excluding Missing Data).

| Characteristic |  | Total<br>(N=19319) | Af Am<br>(N=3766) | Eu Am<br>(N=15553) |
| --- | --- | --- | --- | --- |
| Age |  | 66.4±6.9 | 62.4±6.6 | 67.4±6.7 |
| Female |  | 9767 (50.6) | 2347 (62.3) | 7420 (47.7) |
| BMI |  | 28.0±5.6 | 30.6±6.5 | 27.4±5.2 |
| Current smoking |  | 1309 (6.8) | 528 (14.0) | 781 (5.0) |
| Hypertension medication |  | 9380 (48.6) | 2446 (64.9) | 6934 (44.6) |
| Cholesterol medication |  | 7257 (37.6) | 1200 (31.9) | 6057 (38.9) |
| Diabetes |  | 2429 (12.6) | 871 (23.1) | 1558 (10.0) |
| Diabetes medication |  | 1880 (9.7) | 675 (17.9) | 1205 (7.7) |
| Parental history of MI |  | 3143 (16.3) | 591 (15.7) | 2552 (16.4) |
| Fish consumption (≥1.5/wk) |  | 9100 (47.1) | 1890 (50.2) | 7210 (46.4) |
| Aspirin use |  | 8816 (45.6) | 1431 (38.0) | 7385 (47.5) |
| Statin use |  | 6790 (35.1) | 1114 (29.6) | 5676 (36.5) |
| Vitamin D supplements |  | 8594 (44.5) | 1096 (29.1) | 7498 (48.2) |
| CVD risk factors | 0 | 5833 (30.2) | 778 (20.7) | 5055 (32.5) |
|  | 1 | 6603 (34.2) | 1315 (34.9) | 5228 (34.0) |
|  | >1 | 6883 (35.6) | 1673 (44.4) | 5210 (33.5) |

**Supplementary Fig. 2:**
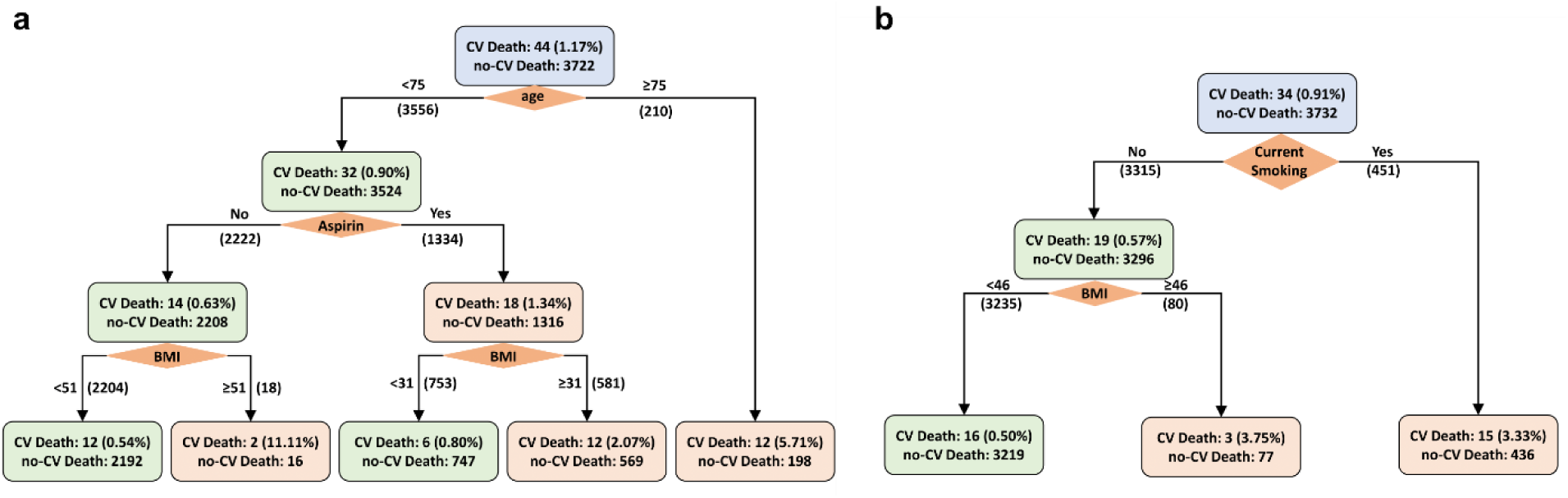
Weighted Decision Tree to Predict Cardiovascular Mortality. a. Tree for AfAm participants after optimal pairs matching, n=3,766, weight ratio = non-diseased : diseased = 3,722:44 = 84.6:1. b. Tree for EuAm participants after optimal pairs matching, n=3,766, weight ratio = non-diseased : diseased = 3,732:3.0 = 109.8:1. age; Age at randomization to VITAL study, years, Aspirin; Baseline aspirin use, BMI; Body mass index at randomization, kg/m2.

**Supplementary Table 2:** Regression Results after Selection of Variables by LASSO with Stroke as the Outcome using AfAm and EuAm Participants after Optimal Pair Matching. Includes interaction terms. Std. Errors, p-values and 95% CI for OR are calculated using non-parametric bootstrap and parametric bootstrap. OR: odds ratio; CI: confidence interval. n-3 HUFA supplementation OR (95% CI) for AfAm subgroup and for EuAm subgroup.

| Variables | Estimate | OR | Non-parametric Bootstrap |  |  | Parametric Bootstrap |  |  |
| --- | --- | --- | --- | --- | --- | --- | --- | --- |
|  |  |  | Std. Error | P-value | 95% CI for OR | Std. Error | P-value | 95% CI for OR |
| (Intercept) | -4.4923 | 0.0112 | 1.9847 | 0.0236 | (0.0002, 0.5476) | 1.9041 | 0.0183 | (0.0003, 0.4675) |
| Vit D Active | -0.5190 | 0.5951 | 0.3762 | 0.1677 | (0.2847, 1.2440) | 0.3967 | 0.1908 | (0.2735, 1.2950) |
| Age | 0.0640 | 1.0661 | 0.0295 | 0.0300 | (1.0062, 1.1296) | 0.0276 | 0.0204 | (1.0100, 1.1254) |
| Current Smoker | 0.6478 | 1.9113 | 0.5720 | 0.2574 | (0.6229, 5.8645) | 0.5095 | 0.2036 | (0.7041, 5.1884) |
| > One risk factor | 0.7410 | 2.0980 | 0.4517 | 0.1009 | (0.8656, 5.0852) | 0.5638 | 0.1887 | (0.6949, 6.3347) |
| n-3 HUFA supplementation x Af Am | 0.4077 | 1.5033 | 0.4377 | 0.3516 | (0.6375, 3.5449)* | 0.4377 | 0.3516 | (0.6724, 3.3612)* |
| n-3 HUFA supplementation x Eu Am | 0.0000 | 1.0000 | 0.0214 | 1.0000 | (0.9589, 1.0428)* | 0.2645 | 1.0000 | (0.5955, 1.6792)* |

**Supplementary Table 3:** Regression Results after Selection of Variables by LASSO with Cardiovascular Death as the Outcome using African American and European American Participants after Optimal Pair Matching. The interaction terms are not selected by model. Std. Errors, p-value and 95% CI for OR are calculated using non-parametric bootstrap and parametric bootstrap. OR: odds ratio; CI: confidence interval.

| Variables | Estimate | OR | Non-parametric Bootstrap |  |  | Parametric Bootstrap |  |  |
| --- | --- | --- | --- | --- | --- | --- | --- | --- |
|  |  |  | Std. Error | P-value | 95% CI for OR | Std. Error | P-value | 95% CI for OR |
| (Intercept) | -5.6986 | 0.0034 | 1.8023 | 0.0016 | (0.0001, 0.1146) | 1.8747 | 0.0024 | (0.0001, 0.1321) |
| Female | -0.6484 | 0.5229 | 0.4225 | 0.1249 | (0.2284, 1.1969) | 0.3875 | 0.0943 | (0.2447, 1.1175) |
| Age | 0.0649 | 1.0671 | 0.0250 | 0.0094 | (1.0160, 1.1206) | 0.0250 | 0.0094 | (1.0160, 1.1206) |
| BMI | 0.0412 | 1.0421 | 0.0246 | 0.0940 | (0.9930, 1.0935) | 0.0265 | 0.1200 | (0.9893, 1.0976) |
| Current Smoker | 1.1492 | 3.1557 | 0.4270 | 0.0071 | (1.3665, 7.2872) | 0.4106 | 0.0051 | (1.4112, 7.0567) |
| Hypertension medication | 0.3457 | 1.4130 | 0.3546 | 0.3296 | (0.7052, 2.8312) | 0.3379 | 0.3063 | (0.7286, 2.7401) |
| Aspirin | 0.3741 | 1.4537 | 0.3085 | 0.2253 | (0.7941, 2.6612) | 0.2951 | 0.2049 | (0.8152, 2.5922) |

